# Mechanistic modeling of social conditions into disease predictions for public health intervention-analyses: application to HIV

**DOI:** 10.1101/2023.03.01.23286591

**Authors:** Chaitra Gopalappa, Amir Khoshegbhal

## Abstract

**Background:** As social and economic conditions are key determinants of HIV, the ‘National HIV/AIDS Strategy (NHAS)’, in addition to care and treatment metrics, aims to address mental health, unemployment, food insecurity, and housing instability, as a strategic plan for the Ending the HIV Epidemic initiative. Mechanistic models of HIV play a key role in identifying cost-effective intervention strategies, however, social conditions are typically not part of the modeling framework. HIV projections are typically simulated by modeling care and sexual behaviors, and transmissions as functions of those behaviors.

**Methods:** We developed a methodological framework, using Markov random field model to estimate joint probability distributions between social conditions and behaviors, to incorporate in a mechanistic model to simulate behaviors as functions of social conditions and HIV transmissions as a function of behaviors. As demonstration, we conducted two numerical applications in a national-level agent-based network model, Progression and Transmission of HIV (PATH 4.0). The first modeled care behavior (using viral suppression as proxy) as a function of depression, neighborhood, housing, poverty, education, insurance, and employment status. The second modeled sexual behaviors (number of partners and condom-use) as functions of employment, housing, poverty, and education status, using exchange sex as a mediator. Both simulated HIV transmissions and disease progression as functions of behaviors. We conducted what-if intervention analyses to estimate the impact of an ideal 100% efficacious intervention strategy.

**Results:** If we intervene on HIV infected persons with the social needs modeled here, such that their care behavior increases to become equal to that among persons who do not have those social needs, the overall viral suppression in persons with diagnosed HIV infection increases from 65.5% to 80% (79% to 83%), resulting in a 10-year cumulative national incidence reduction of 29% (20% to 41%). If we address the social needs modeled here among persons who exchange sex, such that their sexual behavior becomes equal to that among those who do not exchange sex, we can expect a 10-year cumulative national incidence reduction of 6% (2.5% to 14%).

**Conclusions:** We developed a methodological framework for modeling social conditions into intervention decision-analytic models, using the limited data to present two demonstrative applications. Routinely monitoring quantitative data on associations between social conditions and HIV risk behaviors, and efficacy of structural interventions can help develop a comprehensive mechanistic model to identify cost-effective intervention combinations and inform public health strategic plans.

## 1. BACKGROUND

Despite advances in pharmaceutical interventions for the prevention of human immunodeficiency virus (HIV), such as antiretroviral therapy (ART) treatment and pre-exposure prophylaxis (PrEP) that can fully prevent transmissions or acquisition [1], [2], HIV continues to be a huge disease and economic burden in the United States. The number of people with HIV (PWH) was estimated to be 1.2 million in 2019 [3], the number of new infections per year was estimated to be about 35,000 in 2019 [3], and the average discounted lifetime HIV-related medical cost per person was estimated to be $420,285 [4].

There is growing evidence that social determinants of health (SDH), i.e., social and economic conditions, are key drivers of behaviors that increase risk of HIV infection, e.g., lower adherence to care, higher number of partners, higher condomless sex, and higher substance abuse among the homeless compared to those stably housed [5]–[8]. Surveillance of the experiences and needs of persons with diagnosed HIV (PWDH) estimates that about 44% had a disability (including physical, mental, and emotional disabilities), 41% were unemployed, 43% had household incomes at or below the federal poverty threshold, and 10% were homeless [9], [10].

In-line with this evidence, the most recent ‘National HIV/AIDS Strategy (NHAS), Federal Implementation Plan for the United States’, 2022-2025, along with continuing to monitor and target HIV diagnoses, care, and treatment, newly added targets for quality-of-life indicators, such as reduction in unmet needs in mental health services, unemployment, food insecurity, and housing [11]. The goal of the NHAS is to reduce new infections by 75% by 2025 and 90% by 2030 [12]. Alongside biomedical and behavioral interventions, structural interventions are among key evidence-based interventions recommended for HIV prevention [13]. Structural interventions could include a range of programs based on individual needs, such as comprehensive sex education, universal condom availability, expanded syringe access for drug users, health care coverage, subsidized housing and food programs, access to mental healthcare, and early childhood academic enrichment programs[14]–[19]. While the costs of these structural interventions can be extrapolated from small cohort studies, its impact on HIV burden is infeasible to estimate through controlled trials.

Dynamic mathematical simulation models of HIV projections play a key role in evaluation of intervention combinations and inform optimal allocation of intervention resources, including in the context of the NHAS[20]–[24]. These models typically estimate HIV projections through simulation of care and sexual behaviors. There is growing awareness for the need to incorporate SDH into the modeling framework, but thus far, models in this area are limited. Broadly, there are two types of SDH-based models, statistical and mechanistic models [25]. Statistical models, such as regression, directly fit a model between health outcomes (HIV) as response variable and all other metrics including social conditions as independent variables [26], and thus, do not consider the behavioral and disease dynamics. Mechanistic models simulate behaviors, such as sexual behaviors, e.g., number of partners, and condom-use, and care behaviors, e.g., HIV-testing, care retention and treatment adherence, to generate the dynamics of transmission and disease progression [27]–[30].

The focus of our work is to first model care and sexual behaviors as functions of social conditions, incorporated into mechanistic (dynamic) models to then simulate HIV transmissions and progression as functions of behaviors. However, the data to model behaviors as functions of social conditions are not directly available in the national context. Data on prevalence of social conditions are reported mostly as marginal distributions, e.g., proportions in poverty, or homeless, but their joint distributions, e.g., proportion of population in poverty and homeless, for different populations are not available. This is a major data gap, considering the interactions between social conditions. Also, data on associations between behaviors and social conditions are mostly univariate, e.g., correlation between condom-use and housing stability or between number of partners and housing stability, but the multivariate associations, e.g., between condom-use, number of partners, housing stability, and poverty, are not available for all populations. Current models in the literature either provide a general framework that assume parameters are independent or that data are available [25], [27], or have mostly focused on specific populations where individual-level data are available for extraction of the joint distributions, e.g., through randomized control trials that focus on selected individual-level metrics [28], individual-level surveys of a specific population [29], and large longitudinal (∼18 years) individual-level datasets [30]. On one hand, it may be infeasible or expensive to generate such data for all populations, and on the other hand, using one nationally representative dataset would ignore the heterogeneity and thus disparities across populations.

In this work we present a methodological approach for modeling SDH into dynamic mechanistic models and demonstrate the significance of such models for national-level HIV analyses. Specifically, we present a generalized statistical method using Markov random fields or undirected graphical models to infer joint distributions. We then apply the method to two numerical examples to model SDH as functions of behaviors in a national-level Progression and Transmission of HIV (PATH 4.0)[31] dynamic model, and conduct numerical what-if intervention analyses as demonstration of potential national-level analyses. The first numerical example focusses on care behavior using HIV viral load suppression (VLS) as a proxy. The second numerical example focusses on two sexual behaviors, condom use and number of partners, and applied to two sub-populations, heterosexual female and men who have sex with men (MSM). These methods can be easily extrapolated to any number of variables and sub-populations, while maintaining the national-level context through a national simulation model such as PATH 4.0.

## 2. METHODS

### 2.1 Problem description

The general framework of mechanistic models is to simulate HIV transmissions as functions of care behaviors (e.g., viral suppression in Fig 1a) and sexual behaviors (e.g., degree, or number of partners in Fig 1a). Our objective is to expand this to first model care and sexual behaviors as functions of social conditions, such as poverty and homelessness (as seen in Fig 1b).

**Fig 1:**
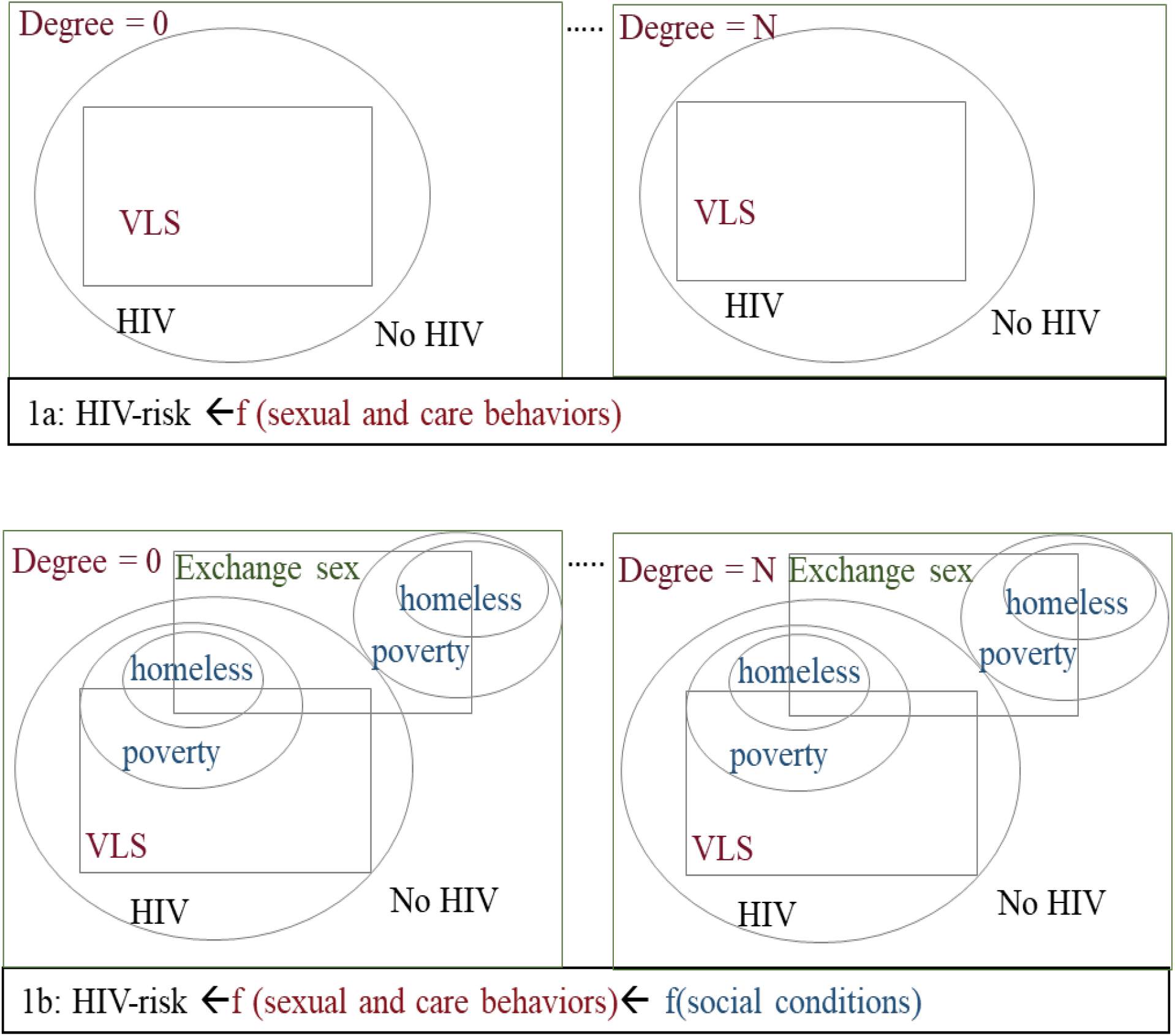
Schematic overview of model framework. 1a(top) Models typically simulate HIV infection as functions of care behaviors (e.g., VLS=> viral suppression from retention-in-care behavior) and sexual behaviors (e.g., degree => number of sexual partners). 1b(bottom) Proposed approach further simulates behaviors as functions of social conditions (e.g., poverty and homelessness).

The data for the marginal distributions of each variable (social conditions or behaviors) are typically available in the literature. Directly sampling from these distributions would assume independence between variables, which is contrary to the evidence of correlations observed in the literature. Examples of correlations related to care and sexual behaviors are presented in Fig 2 and Fig 3, respectively. Each link/edge in Fig 2 and Fig 3 depict observations of significant associations or correlations between the variables they connect, that we quantitatively represent through relative risk (RR). For example, as seen in equation for pairwise variables [VLS, housing] in Fig 2, RR is the probability of no-VLS (i.e., not in care or treatment) among persons who are homeless divided by the probability of no-VLS among persons who are not homeless, a value greater than 1 suggesting correlations between VLS and housing. Several such associations are observed in the literature either directly between any two social conditions, or a social condition and a behavior, or through intermediary variables (mediators) such as depression in Fig 2 or exchange sex in Fig 3, that are relevant to include in the model to highlight other healthcare or social needs. However, the estimates in the literature, which are primarily observational studies, are restricted to pairwise associations, the associations between all variables are not fully known and typically challenging to estimate through observational studies alone.

**Fig 2:**
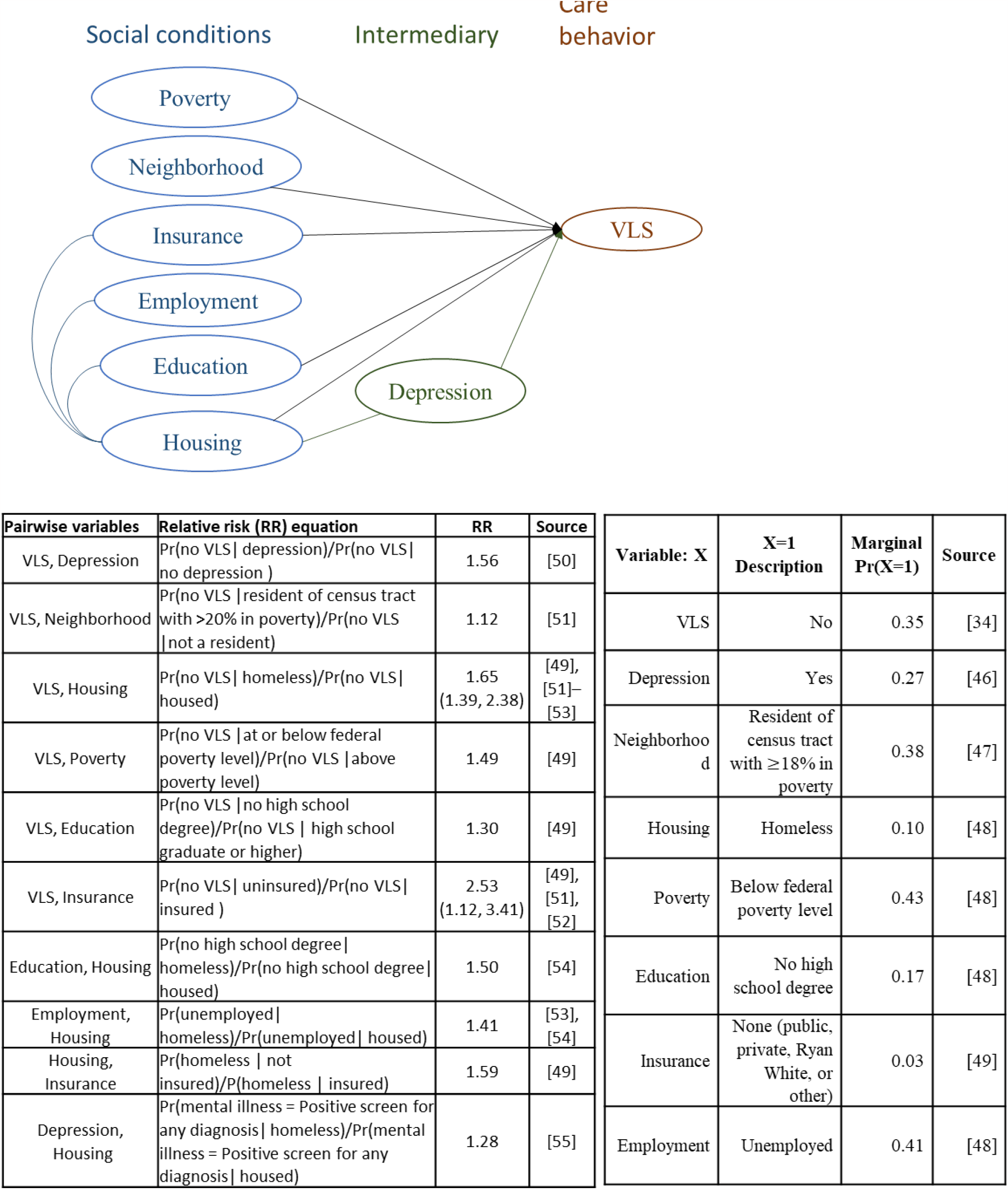
(clockwise from top) Illustration of associations between social conditions and care behavior, marginal distributions, and relative risk between pairwise variables. Note: all variables are binary.

**Fig 3:**
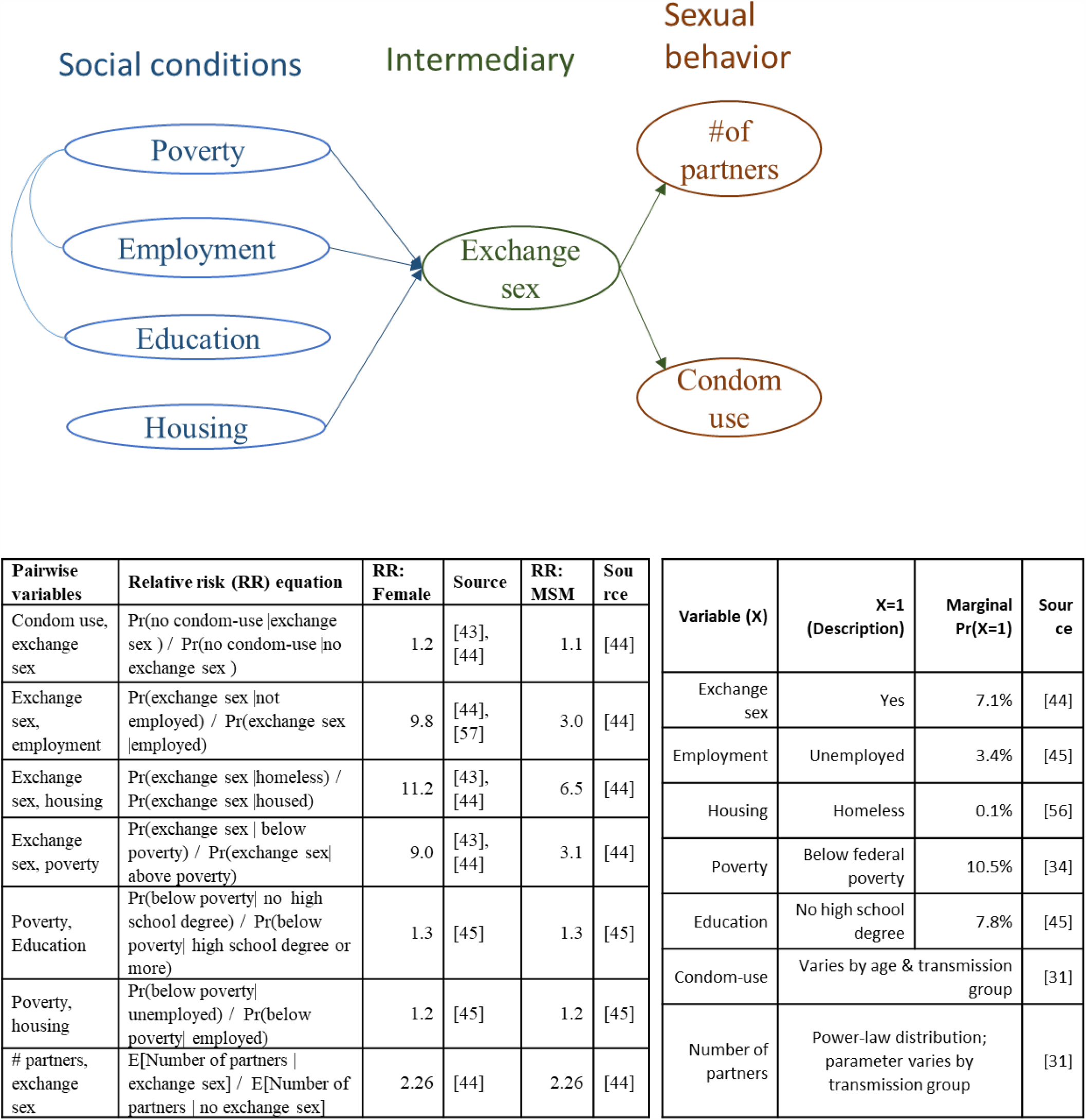
(clockwise from top) Illustration of associations between social conditions and sexual behavior, marginal distributions, and relative risk between pairwise variables. Note: all variables except number of partners are binary.

The problem is to thus estimate the joint associations between all variables. For the problem in Fig 2, this will be to estimate the joint probability, Pr(*VLS, D, N, H, P, E, I, W*) where, D=depression, N=neighborhood, H=housing, P=poverty, E=education, I=insurance, and W=employment, each feature a binary variable taking value of 0 to represent a good status (e.g., housed) or 1 to represent a socially disadvantaged status (e.g., homeless). The joint probabilities would provide the flexibility to simulate variables of interest, e.g., to first simulate the SDH status of an individual by sampling from Pr(*D, N, H, P, E, I, W*) and then simulating retention-in-care behavior (using VLS as proxy here) as a function of SDH status through use of a conditional distribution, estimated as Pr(*VLS*|*D, N, H, P, E, I, W*) = Pr(*VLS, D, N, H, P, E, I, W*)Pr (*VLS*). Note that, here we are assuming causality between care behavior and social conditions, but not between social conditions. Similarly, the problem corresponding to Fig 3 is to estimate the joint distribution of all variables, so that, in a simulation model, we can add SDH status by sampling from a joint distribution of social conditions, simulate exchange sex as a function of SDH status (assuming causality), and simulate number of partners and condom use as functions of exchange sex (assuming causality), subsequently simulating HIV transmissions as functions of behaviors. We assume all variables in Fig 2 and Fig 3 are binary except for number of partners which takes integer values between 0 and the maximum number of partners.

### 2.2 Markov random field (MRF) model

Problem formulation: The problem can be represented as an undirected graphical model, also known as Markov random field model (MRF) [32], with each variable represented by a node in a graph and the data associations between variables represented by an edge connecting the nodes. This representation will be similar to Fig 2 and Fig 3, except that the edges are undirected, to estimate the joints without assuming causality (although in the intervention analyses we will simulate behaviors casual to social conditions, but assume no causality between any two social conditions). Specifically, the joint probability vector (***y***) can be estimated by solving for the parameter vector ***θ*** in the following equation,

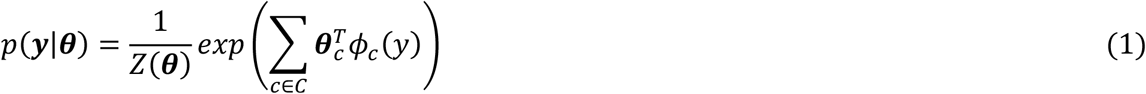

where, *ϕ*_*c*_(*y*) is a feature vector corresponding to each feature *c* (here, we consider each edge/link in the graph as a feature), *C* is the set of all features, and *Z*(***θ***) is the normalizing constant.

The equation in (1) can be derived using maximum entropy. The objective of a maximum entropy model is to pick the distribution *p*(***y***) with maximum entropy (− Σ_*y*_ *p*(*y*) log *p*(*y*)) (note that it would be the one closest to uniform) subject to the constraints that the moments of the distribution match the empirical moments of the specific feature vectors *ϕ*_*c*_ (*y*), ∀*c* ∈ *C*, included in the objective function through vector Lagrange multipliers (***θ***_*c*_, ∀*c* ∈ *C*). If there is a solution which is a distribution (i.e., *p*(*y*) ≥ 0, and Σ_*y*_ *p*(*y*) = 1) and is the maximal entropy solution, then the unique distribution is of the form given in (1).

As the MRF in (1) belongs to the exponential family, its scaled log-likelihood (in (2) below) is concave on ***θ*** and thus the joint probability can be estimated using gradient descent method, by solving for the value of ***θ*** that provides a zero gradient [32].

The log-likelihood of (1) can be written as

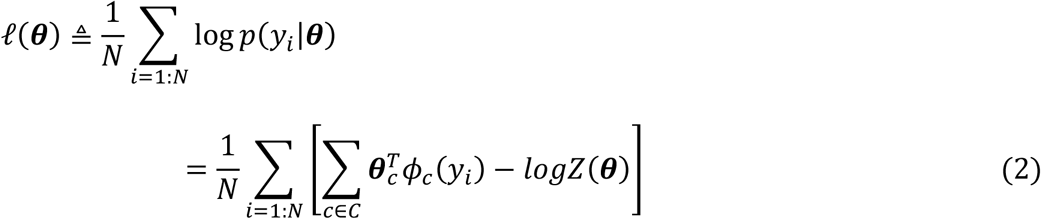

The gradient of the log-likelihood in (2) can be written as the expected feature vector according to the empirical distribution minus the model’s expectation of the feature vector as follows,

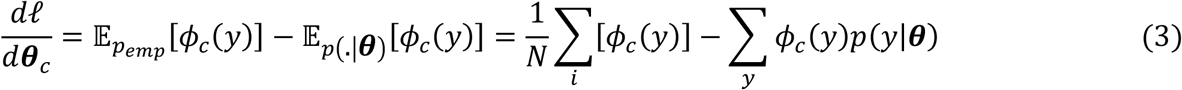

Thus, the joint probability can be estimated by solving for ***θ*** where the empirical distribution (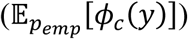) is equal to the model’s expectation of the feature vector (Σ_*y*_ *ϕ*_*c*_ (*y*)*p*(*y*|***θ***)). We apply gradient descent method to solve for ***θ***, i.e., by starting with an arbitrary value of ***θ***, we can find its optimal value by iterating through the equation,

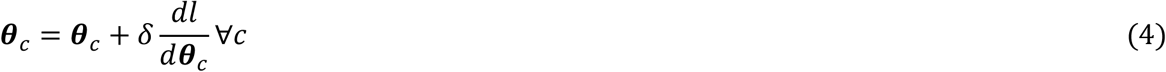

where *δ* is the step-size, until 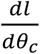 converges to zero. Estimation of the two components in (3) are discussed under numerical applications I and II, and correspond to the configurations in Fig 2 and Fig 3, respectively.

### 2.3 Numerical Application I: Care Behavioral Model

#### 2.3.1 Overview of care behavioral metric and social conditions

We chose viral load suppression (VLS) as a proxy for care behavioral metric. Though the care continuum stages for persons diagnosed with HIV infection and linked to care include receipt of care, retention-in-care (measured as one or more CD4 count or viral load tests), and VLS (when viral load <200 copies/mL) [33], the number of studies evaluating each of these stages separately as related to social conditions is limited. The metrics used in the literature to represent each of these stages also vary, and include prescribed ART, taking ART, one or more lab tests, or VLS. Further, VLS is achieved through consistent-use of ART, and thus, retention-in-care behavior leads to viral suppression. Therefore, we assumed VLS as a proxy for overall care behavior, using its marginal distribution from the U.S. National HIV Surveillance System[34] (Fig 2). For the relative risk metrics between social conditions and care behavior, we gathered data for any of the metrics noted above and conducted a sensitivity analyses using the range of values (variables, metrics, and data sources are in Fig 2).

The metrics related to social conditions were also not comprehensive to maintain consistency in the variables we used, e.g., as we did not have data for associations between Depression and Housing, we assumed it would be equal to associations between Mental health and Housing. Most metrics also had only one study. For metrics with more than one study, we used the median value as the basecase and the minimum and maximum values for a sensitivity analysis to generate a range in output results. Variables, metrics, and data sources are presented in Fig 2.

#### 2.3.2 Estimating joint distribution using MRF: care behavioral model

Corresponding to Fig 2, we have eight variables (*K* = 8), one care behavior indicator (VLS), one intermediary (depression), and six social conditions, thus *y* is a vector of dimension *K* (*y* = [*VLS, D, N, H, P, E, I, W*]). We assume that each edge in Fig 2 is a feature, and thus the model in (3) will only contain pairwise features, i.e.,

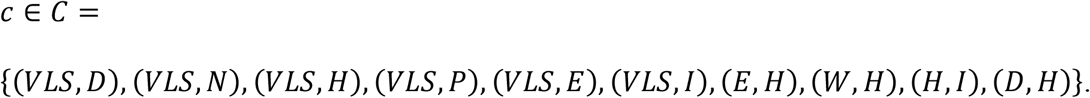

We assume that all variables are binary, and thus, for each edge, we have a feature vector of size 2^2^, i.e.,

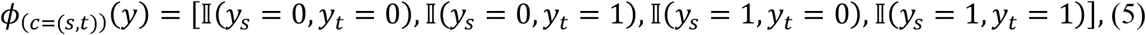

where, 𝕀 is an indicator function, i.e., 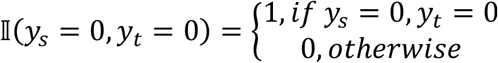. For example, *ϕ*_(*c*=(*W,H*))_(*y* = [1,0,0, *H* = 0,1,0,1, *W* = 1]) = [0,0,1,0].

Thus, for any given ***θ***, the second component in (3), 𝔼_*p*(.|***θ***)_ [*ϕ*_*c*_ (*y*)], can be calculated for every value of *y* using *ϕ*_*c*_ (*y*) from (5) and *p*(*y*|***θ***) from (1). The first component in (3), 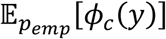, is the number of times that feature appears in the data, and thus, in the context here, it will be equivalent to the pairwise joint probabilities, which we estimated using literature data.

Specifically, we use the relative risk (RR) values from the literature, which can originate from smaller samples, and the marginal distributions corresponding to the population of interest (nationally representative in our numerical examples), to calculate the joint probabilities. Below is an example taking feature *c* = (*S*_1_, *S*_2_).

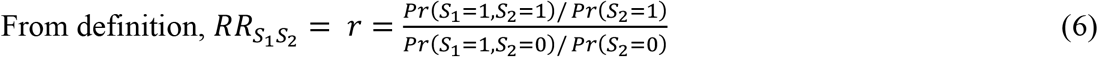

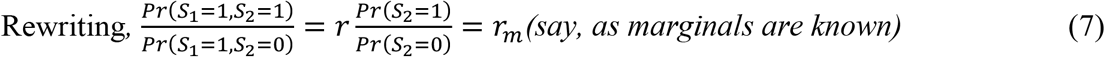

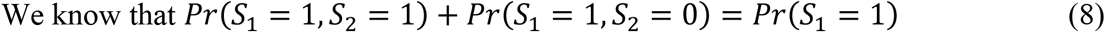

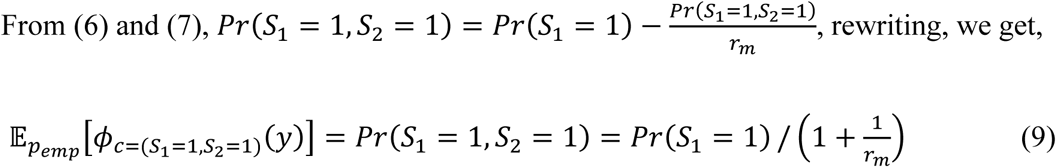

As the marginal distributions and relative risk values (Fig 2) are known (i.e., the R.H.S. of (9) can be calculated using literature data), we can estimate *Pr*(*S*_1_ = 1, *S*_2_ = 1) from (9), and consequently, through simple probability rules, also estimate *Pr*(*S*_1_ = 1, *S*_2_ = 0), *Pr*(*S*_1_ = 0, *S*_2_ = 1), and *Pr*(*S*_1_ = 0, *S*_2_ = 0), and thus all components corresponding to feature *c* = (*S*_1_, *S*_2_). Note that, though in some cases the literature studies may directly present the pairwise joint probabilities (*Pr*(*S*_1_, *S*_2_)), they may be estimated from smaller samples and thus may not be representative of the population modeled in the simulation. This method of using the relative risk with more representative marginal distributions avoids sample bias, but with the caveat that it assumes that the RR observed in small samples are representative of the larger population.

#### 2.3.3 What-if impact analyses: care behavioral model

Using the joint distributions, we can calculate the joint probabilities of SDH (Pr (*SDH*)) and conditional probabilities Pr (*VLS*|*SDH*) for any combination of SDH, which can be used in a mechanistic simulation to add SDH status and simulate care behavior as a function of SDH status. As demonstration of potential significance of such a simulation model, we conducted the following what-if analyses. From literature data, we know that the *Pr*(*VLS* = 1|*SDH* = ***a***_*n*_) > *Pr*(*VLS* = 1|*SDH* = **0**_*n*_), ∀***a***_*n*_ ≠ **0**_*n*_, where value of 0 is a good status and 1 is a bad status, *SDH* is some *n*-combination of social conditions, **0**_*n*_ is a vector of *n*-zeros indicating a good status for all *n*-social conditions, and ***a***_*n*_ is a binary vector of *n*-values with at least one 1 indicating at least one socially disadvantaged condition. We evaluated the impact of *Pr*(*VLS* = 1|*SDH* = ***a***_*n*_) → *Pr*(*VLS* = 1|*SDH* = **0**_*n*_), i.e., suppose we are able to intervene on those people with the social needs, such that their chance of VLS increases to become equal to that among those with no social needs, what would be the corresponding impact? To note here that transmission rate from persons with VLS is close to zero, and thus, in addition to having therapeutic benefits for the HIV infected person, VLS will also reduce incidence (new infections). Therefore, we evaluated two metrics of impact, taking all *n* = 7 social conditions from Fig 2. In the first, as a metric for improvement in care among PWDH, we calculated the expected maximum VLS level, i.e., 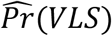 when setting 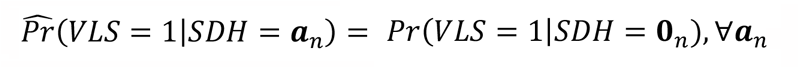. In the second, as a metric of population-level impact, we estimated the cumulative reduction in new infections over a 10-year period, by simulating two scenarios, a baseline scenario using status-quo marginal distributions (*Pr*(*VLS*)) and an intervention scenario using 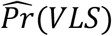. These analyses were conducted in the PATH 4.0, which is a national simulation model of HIV in the United States and validated to be representative of the HIV epidemic over the period 2006 to 2017 (see brief overview below). We modeled SDH and VLS such that the marginal distribution for VLS, for the period 2018 to 2027, was kept at the 2017 value (*Pr*(*VLS*)) in the baseline scenario and at the intervention value 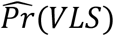 in the intervention scenario. We simulated 30 runs and present the mean and range across the runs.

#### 2.3.4 Overview of PATH 4.0 model

PATH 4.0 (Progression and Transmission of HIV) is a comprehensive simulation model of HIV in the United States developed using a new agent-based evolving network modeling (ABENM) simulation technique [35]. The main concept of ABENM is to simulate persons infected with HIV and their immediate contacts (all sexual partners a person will have over their lifetime) as individual agents and all other persons using a compartmental model. An Evolving Contact Network Algorithm (ECNA) maintains the network dynamics between the agents and the compartment, transitioning persons from the compartment to agents as the disease spreads. Considering the low prevalence of HIV in the U.S. (0.4%), a national network model of HIV is computationally impractical. ABENM addresses this gap by using a hybrid simulation technique while maintaining the network features through ECNA. The model was validated for the period 2006 to 2017 by calibrating to 2006 data, simulating the epidemic from 2006 to 2017 in monthly-time steps, and comparing simulated estimates for multiple features, including population-level epidemic features and HIV-network features, the latter using molecular clusters, using data from surveillance and surveys such as the National HIV Surveillance System (NHSS), the Medical Monitoring Project (MMP), the HIV Outpatient Study (HOPS), the National HIV Behavioral Surveillance (NHBS), the National Survey for Family Growth (NSFG), and the National Survey for Sexual Health and Behavior (NSSHB)[36]–[41]. Details of the model and validation are presented elsewhere [31]. Though we restricted this demonstration case study to the national-level, PATH 4.0 provides a suitable national-level framework for modeling social conditions and their heterogeneity across populations, while maintaining the overall network dynamics.

As PATH 4.0 tracks HIV-infected person as agents, we added social conditions to each agent using the joint distributions, and modeled VLS conditional on SDH, simulating for the period 2018 to 2027, maintaining the 2017 marginals (*Pr*(*VLS*)) in the baseline scenario and the intervention marginals *Pr*(*VLS*) in the intervention scenario. As PATH 4.0 simulates HIV transmissions as a function of care behaviors (VLS) and sexual behaviors, we can measure the impact of the intervention by the differences in the number of new infections between the two scenarios.

### 2.4 Numerical Application II: Sexual Behavioral Model

#### 2.4.1 Overview of sexual behavioral metrics and social conditions

We focused the analysis on persons who exchange sex (though we model the full US population in PATH 4.0), as it was the main mediator associating sexual behavior with social conditions for which data were available (Fig 3). The main data source was the National HIV Behavioral Surveillance system [42], a comprehensive surveillance of behavioral risk factors conducted in three populations, men who have sex with men (MSM), injecting drug users, and high-risk heterosexual. Generally, persons reporting receiving ‘things like money or drugs in exchange for sex’ were categorized as persons who exchange sex. For heterosexuals, as data related to exchange sex was restricted to persons in low socio-economic neighborhoods [43](numerator in relative risk estimations in Fig 3), for the data on exchange sex among persons without the social condition (denominator of relative risk), we made a simplifying assumption of using the data from MSM[44]. As PATH only models sexual transmission of HIV, we excluded injecting drug users.

All variables, data on marginals and relative risk, and data sources are presented in Fig 3. The relative risk between any two social conditions *X* and *Y*, specifically Poverty and Education, and Poverty and Employment in Fig 3, were not directly available in the literature. We derived them using census-tract data [45] on the marginals (*Pr*(*X*), *Pr*(*Y*)), solving for the pairwise joint distribution (*Pr*(*X, Y*)) using Pearson correlation coefficient 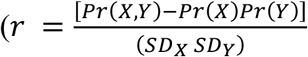; and standard deviation (SD) estimated from the census-tract data), and subsequently using the joint to calculate the conditionals in the RR equation in Fig 3.

We conducted this analysis independent of the care behavioral model in Numerical analyses I. This is because, though we modeled behaviors as a function of social conditions, its association was only through exchange sex, and therefore, the resulting distributions of social conditions among persons with HIV infection would not be fully representative of reality.

Though our analyses is restricted to a small population, it provides a concrete example for a demonstration case study of the proposed methodological framework for modeling social conditions as part of disease prediction modeling, and suitable analyses from such a model.

#### 2.4.2 Estimating joint distribution using MRF: sexual behavioral model

Corresponding to Fig 3, we have seven variables (*K* = 7), two sexual behavioral variables (condom use, and number of partners), one intermediary variable (exchange sex), and four social conditions, thus *y* is a vector of dimension *K*. All variables except number of partners are binary, and thus the joint distribution between the six variables (exchange sex, condom-use, housing, education, employment, and poverty) can be estimated similar to that in care behavior. For simulating all variables in PATH, we take a sequential approach as follows, which is an approach most suitable for the network context in PATH (however, note that, once we have the joint distributions, there is flexibility in determining the sequence of modeling that is most suited to an application).

#### 2.4.3 Parameterization of SDH in PATH 4.0 simulation

In the PATH 4.0 model, sexual partnership networks follow a scale-network, i.e., the life-time number of partners per person (degree *D*) follows a power-law distribution Pr(*D* = *d*) ∼*d*^−*λ*^, *λ* is the power-law parameter. For every person in the simulation, the model determines degree (life-time number of partners) by using a machine learning algorithm that was trained to maintain this overall scale-free network structure, and partnerships of each person are activated and deactivated over time/age using a set of optimization algorithms[31]. Further, condom-use changes as persons age[31]. Thus, to ensure the network and behavioral dynamics are maintained, we take a sequential approach to model parameterization of SDH as follows. First, assign degree (lifetime partners) using typical PATH 4.0 method, as summarized above. Second, assign exchange sex (*B*) conditional on degree (*D*), using

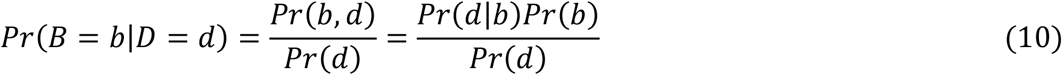

The marginals *Pr*(*D*) and *Pr*(*B*) are available in the literature (Fig 3). Assuming *Pr*(*D* = *d*|*B* = 1) and *Pr*(*D* = *d*|*B* = 0) also follow scale-free property with parameters *λ*_*B*=1_ and *λ*_*B*=0_, respectively, the conditional distributions were estimated by numerically solving, through trial and error, for the values *λ*_*B*=1_ and *λ*_*B*=0_, that provided best fit to the following,

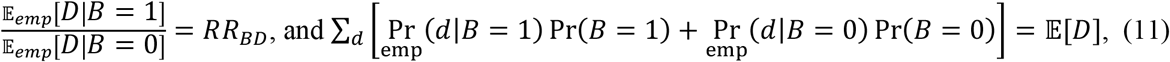

where, 𝔼_*rmp*_[*D*|*B* = *b*] is the empirical expected degree and 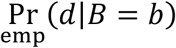 the empirical conditional probability calculated for a specific *λ*_*B*=*b*_, *RR*_*BD*_ is the literature value for expected degree among persons who exchange sex relative to expected degree among persons who do not exchange sex (Fig 3), and 𝔼[*D*] is the expected degree used in the development of PATH 4.0 model[31].

Third, assign SDH status (H=housing, P=poverty, W=employment, E=education) using

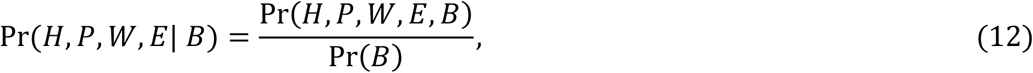

where, Pr(*H, P, W, E, B*) can be estimated using (1) through (4), and Pr(*B*) is known from the literature (Fig 3).

Finally, assign condom-use (*C*) using,

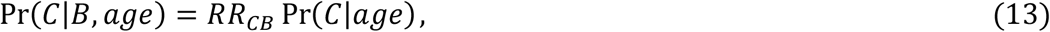

where, *RR*_*CB*_ is the literature value of relative risk of no condom-use among persons who exchange sex relative to those who do not (Fig 3), and the Pr(*C*|*age*) are the distributions used in the PATH model. Note that, limited by data, this method does not model longitudinal changes in individual-level behaviors of exchange sex. In the absence of such data, the method noted above of modeling lifetime number of partners accounts for the overall differences in life-time number of partners between persons who have ‘ever’ exchanged sex sometime during their lifetime and those who have ‘never’, and the assumption of power law distribution accounts for the differences, in number of times and partners, among those who have ever exchanged sex. The caveat being the validity of these assumptions cannot be directly verified, we only conducted an overall validation on HIV cases among persons who exchange sex, as discussed in results.

#### 2.4.4 What-if impact analyses: sexual behavioral model

As per literature data, we know that 𝔼(*D*|*exchange sex* = *yes*) > 𝔼(*D|exchange sex* = *no*), and Pr(*condom use* = *no|exchange sex* = *yes*) >

Pr(*condom use* = *no|exchange sex* = *no*). We evaluated the impact of 𝔼(*D|exchange sex* = *yes*) → 𝔼(*D|exchange sex* = *no*), and Pr(*condom use* = *no|exchange sex* = *yes*) → Pr(*condom use* = *no|exchange sex* = *no*). That is, suppose we are able to address the social needs of persons who exchange sex, such that their sexual behavior becomes similar to those who do not exchange sex, what is the corresponding impact? We measured impact by estimating reduction in cumulative new infections over a 10-year period (2017 to 2028) by simulating two scenarios, a baseline scenario that assumed degree and condom-use distributions as in original data, and a second intervention scenario that assumed degree and condom-use distributions among persons who exchange sex are equal to that among persons who do not exchange sex. We simulated 30 runs and present the mean and range across the runs.

## 3. RESULTS

### 3.1 Numerical application I: Care behavioral model

From the estimated joint distribution, presented in Fig 4 as a density function on the number of social conditions, about 12% of all people with diagnosed HIV (PWDH) have 0 social conditions, ∼15% of PWDH with VLS have 0 social conditions, and ∼7% of PWDH with no-VLS have 0 social conditions. That is, we can infer that most PWDH have at least one social condition, and PWDH with VLS have fewer social conditions than PWDH without VLS. From Fig 4 we can also infer that most PWDH (∼63%) have 1 or 2 social conditions, and fewer (∼25%) have >2 social conditions with proportion having *k* conditions decreasing as *k* increases.

**Fig 4:**
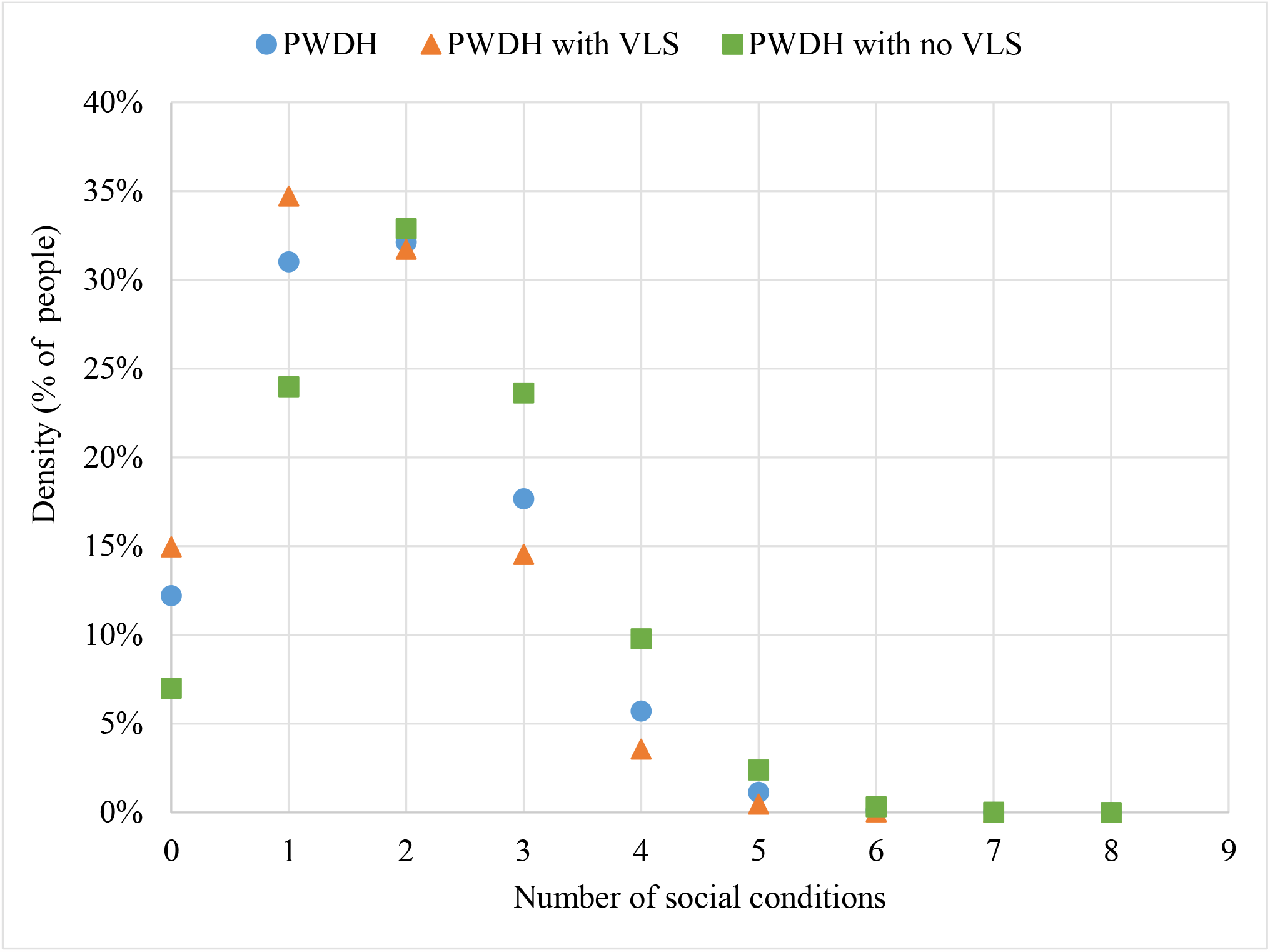
Probability density function (% of people) for the number of social conditions in the population (for the social conditions modeled here -see Fig 2). PWDH: people with diagnosed HIV; VLS: viral load suppression.

The relative risk metrics (i.e., the correlations) between all pairwise variables are presented in Table 1. The values in shaded cells are the inputs used for the development of the MRF model, we verify that the model reproduces the inputs. All other values are an outcome of the MRF model. The results highlight the significance of data gaps. For example, the relative risk between unemployment and poverty is 1, suggesting that they are independent variables, which is contradictory to intuition. To recollect, the principles of MRF model is to find a distribution with maximum entropy subject to constraints (features)(here, the shaded values in Table 1). The distribution closest to uniform, i.e., assumption of independence between variables, has the maximum entropy, and thus, any deviations from independence are a manifest of the applied constraints. In the case of Employment and Poverty, an assumption of independence still provided a good fit to all constraints (the value in shaded cells) thus resulting in a RR value of 1. In some other cases, e.g., Insurance and Depression, independence assumption did not give a good fit to all constraints, thus resulting in a value of RR >1. Therefore, more the data, even if not direct associations, more accurate the estimations.

**Table 1:** Relative risk (RR)* estimates for all pairwise variables

|  | **B→** | VLS | Depression | Neighborhood | Housing | Poverty | Education | Insurance | Employment |
| --- | --- | --- | --- | --- | --- | --- | --- | --- | --- |
| **A** | **Description (B=1)\*→** | No | Yes | Resident of census tract with ≥18% in poverty | Homeless | Below federal poverty level | No high school degree | None (public, private, or other) | Unemployed |
| VLS |  | - | 1.56 | 1.12 | 1.65 | 1.49 | 1.30 | 2.52 | 1.04 |
| Depression |  | 1.67 | - | 1.02 | 1.28 | 1.08 | 1.06 | 1.28 | 1.02 |
| Neighborhood |  | 1.15 | 1.02 | - | 1.03 | 1.02 | 1.01 | 1.05 | 1.00 |
| Housing |  | 2.20 | 1.37 | 1.03 | - | 1.13 | 1.58 | 1.60 | 1.80 |
| Poverty |  | 1.41 | 1.06 | 1.01 | 1.08 | - | 1.03 | 1.18 | 1.00 |
| Education |  | 1.41 | 1.07 | 1.01 | 1.51 | 1.05 | - | 1.19 | 1.03 |
| Insurance |  | 9.66 | 1.41 | 1.09 | 1.67 | 1.35 | 1.23 | - | 1.04 |
| Employment |  | 1.03 | 1.01 | 1.00 | 1.41 | 1.00 | 1.02 | 1.02 | - |

\*Relative risk (RR) of A with respect to B= $\Pr(A=1|B=1)/\Pr(A=1|B=0)$ ; 0 representing good social status and 1 representing socially disadvantaged status.

The baseline %VLS among PWDH in the United States was about 65.5% (Fig 5). If an intervention strategy successfully improves ‘all’ the social needs of people, such that their VLS increases to become equal to the VLS among those with no SDH, we can achieve upto 80% (79% to 83%) VLS level (Fig 5), the range resulting from the sensitivity analyses using lower and upper bound of RR metrics when more than one data source was available (Fig 2). The change in %VLS levels if the ‘individual’ social conditions were intervened varied across the social conditions (Fig 5), influenced by the proportion of PWDH with the condition (marginals) and the relative risk (RR) metrics used in model development (Fig 2). For example, if an intervention addresses depression, which was prevalent among 27% of PWDH, we can achieve upto 70% VLS level, whereas, if an intervention addresses neighborhood poverty, which was prevalent among 38% of PWDH, we can achieve only upto 67% VLS (Fig 5). These results are as expected because the corresponding relative risk values used in model development were 1.56 for no VLS given depression and 1.12 for no VLS given neighborhood (Fig 2). The results also highlight the significance of any data gaps. For example, although unemployment was prevalent among 41% of PWDH, if an intervention addresses unemployment we can achieve only upto 66% VLS level (Fig 5), as the relative risk related to VLS and Employment was only 1.04, an output from the MRF model as there was no data to include it as a constraint in the MRF. As noted earlier, the distribution closest to independence, subject to constraints, has the maximum entropy, and thus the RR between VLS and Employment is the result of indirect constraints (between VLS and Housing and between Housing and Employment (Fig 2)).

**Fig 5:**
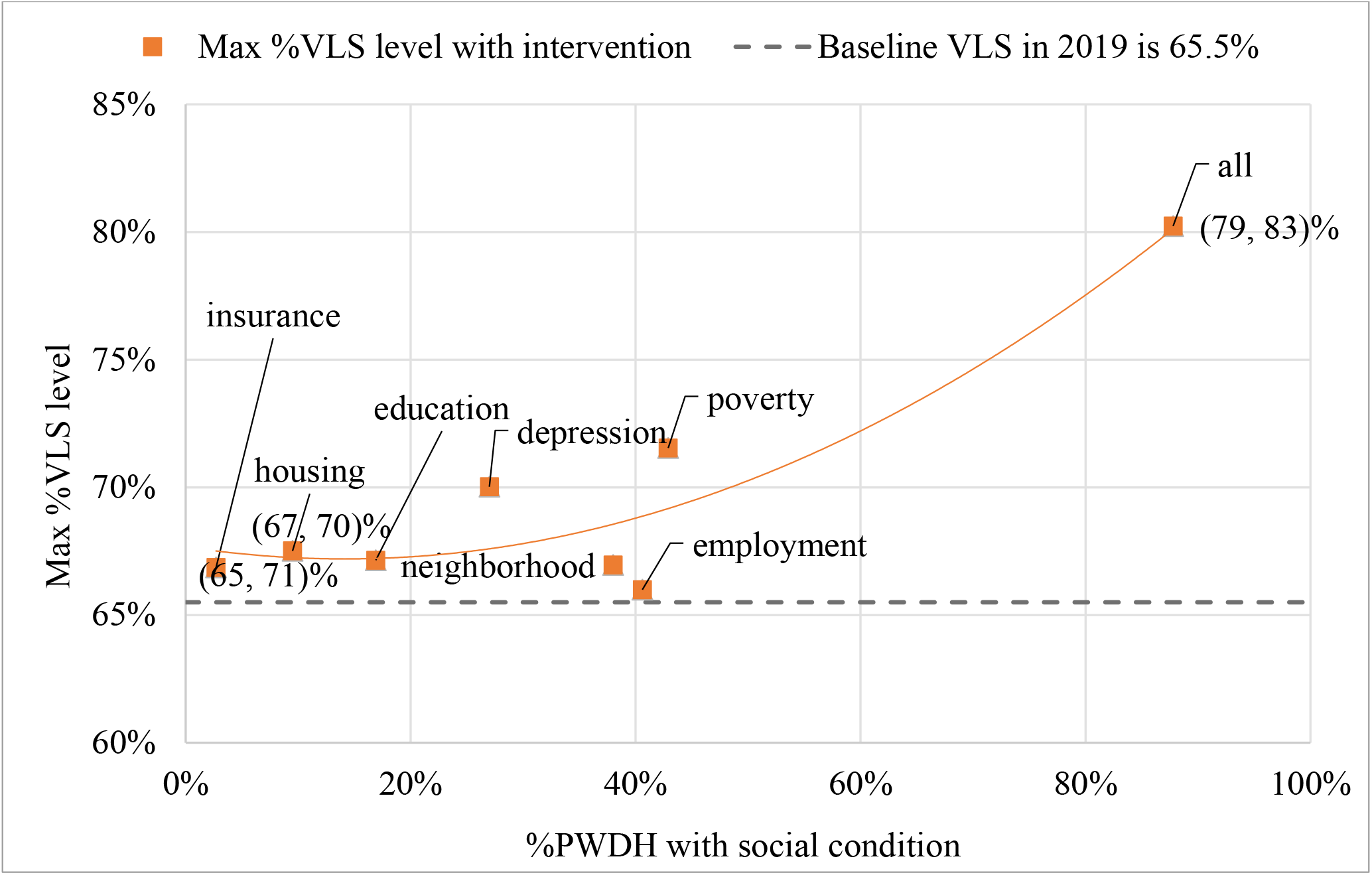
Maximum %VLS level if an intervention strategy successfully improves VLS levels of PWDH with the social condition to equal that of PWDH without the social condition; PWDH: persons with diagnosed HIV; VLS: HIV viral load suppression. Range denote outcome from sensitivity analyses, conducted only for metrics with multiple data sources.

If an intervention strategy successfully improves ‘all’ the social needs of people, i.e., we achieve 80% VLS level (Fig 5), then we can expect a 10-year cumulative incidence reduction of 29% (20% to 41%), the average and range is across 30 simulation runs.

### 3.2 Numerical analyses II: Sexual behavioral model

As expected, the joint distribution, presented as a density function of the number of social conditions in Fig 6, suggests that the proportion of people with 0 social conditions is higher among persons who did not engage in exchange sex compared to those who did (Fig 6). While about 26% (and 84%) of heterosexual women with exchange sex (and no exchange sex) had 0 SDH, about 61% (and 81%) of MSM with exchange sex (and no exchange sex) had 0 SDH. Though the marginal distributions for exchange sex was assumed the same for women and MSM, the differences can be expected from the relative risk values between exchange sex and social conditions, used as inputs for the MRF model, which were significantly higher for women than MSM (*Fig 3*).

**Fig 6:**
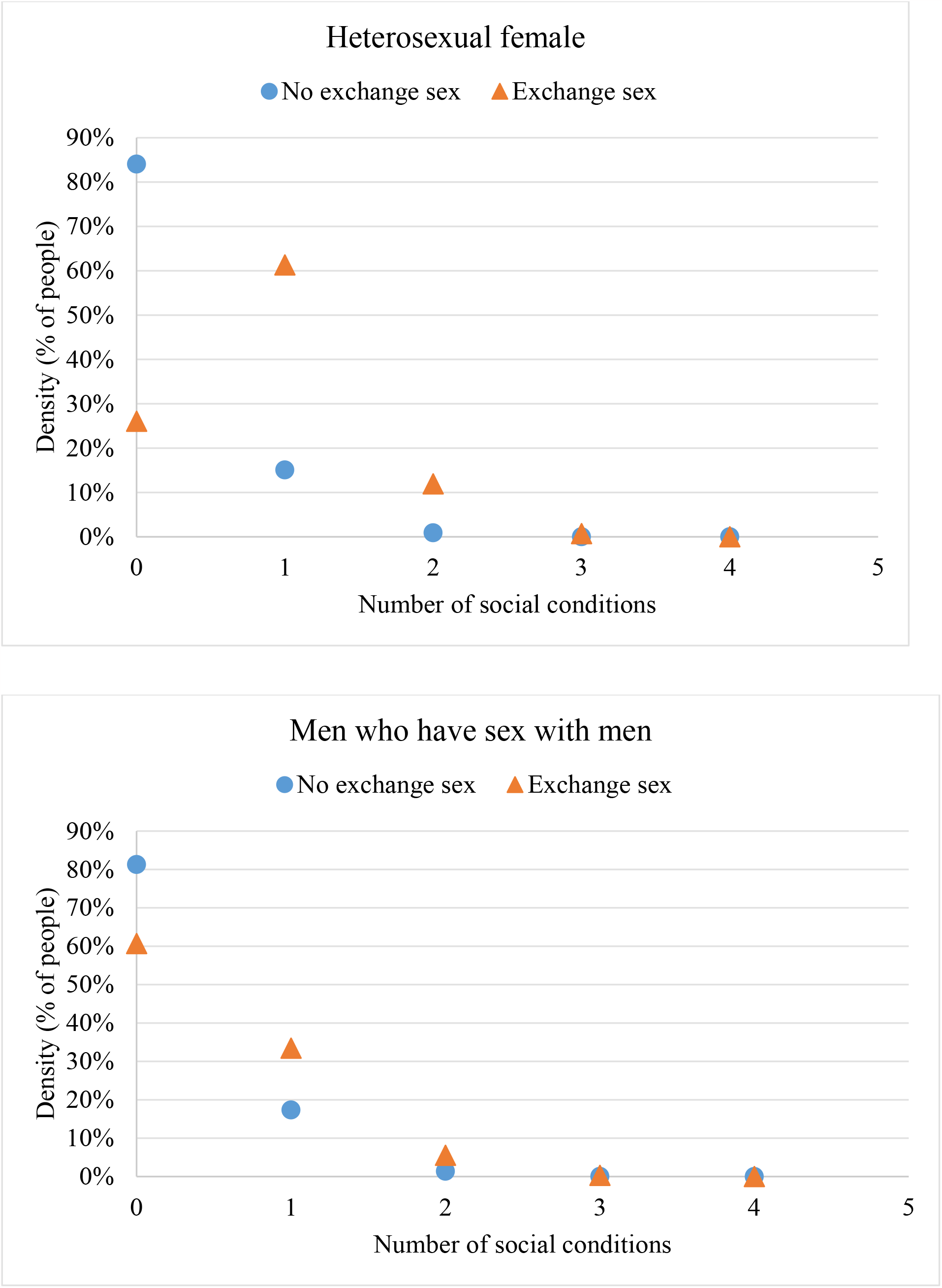
Probability density function (% of people) for the number of social conditions in the population (for the conditions modeled here see Fig 3), split by exchange sex status.

In this numerical example, the model inputs were distributions among the general population and were included in the simulation as such, i.e., each person in the general population was assigned an SDH status drawing from the joint distribution, and their sexual behaviors were modeled as a function of their SDH status. Further, as in typical mechanistic models, HIV transmissions were modeled as a function of behaviors. Therefore, the distributions of SDH among HIV infected persons are an outcome of the dynamics of the simulation model. Thus, for model validation, we compare model estimated results with literature estimates for metrics among PWH, where available (Table 2a). To note here that the literature estimates were among all transmission groups, including people who inject drugs, however, in the PATH 4.0 model we only simulate sexual transmission groups. As the literature study did not present HIV cases by transmission group but only exchange sex status, we made a simplifying assumption of excluding the number of persons who inject drugs from all numbers (HIV, exchange sex, and no exchange sex), and thus the modified literature values underestimate sexually transmitted HIV prevalence. Considering these modifications, we verify that the model estimates are close to the literature estimates, and though not within the range, are in the expected direction (i.e., as literature values for Pr(HIV|ExSex) and Pr(HIV|No ExSex) are an underestimation, they are lower than model estimates, and as literature values for Pr(ExSex| HIV) is an overestimation, they are higher than model estimates) (Table 2a).

**Table 2a:**
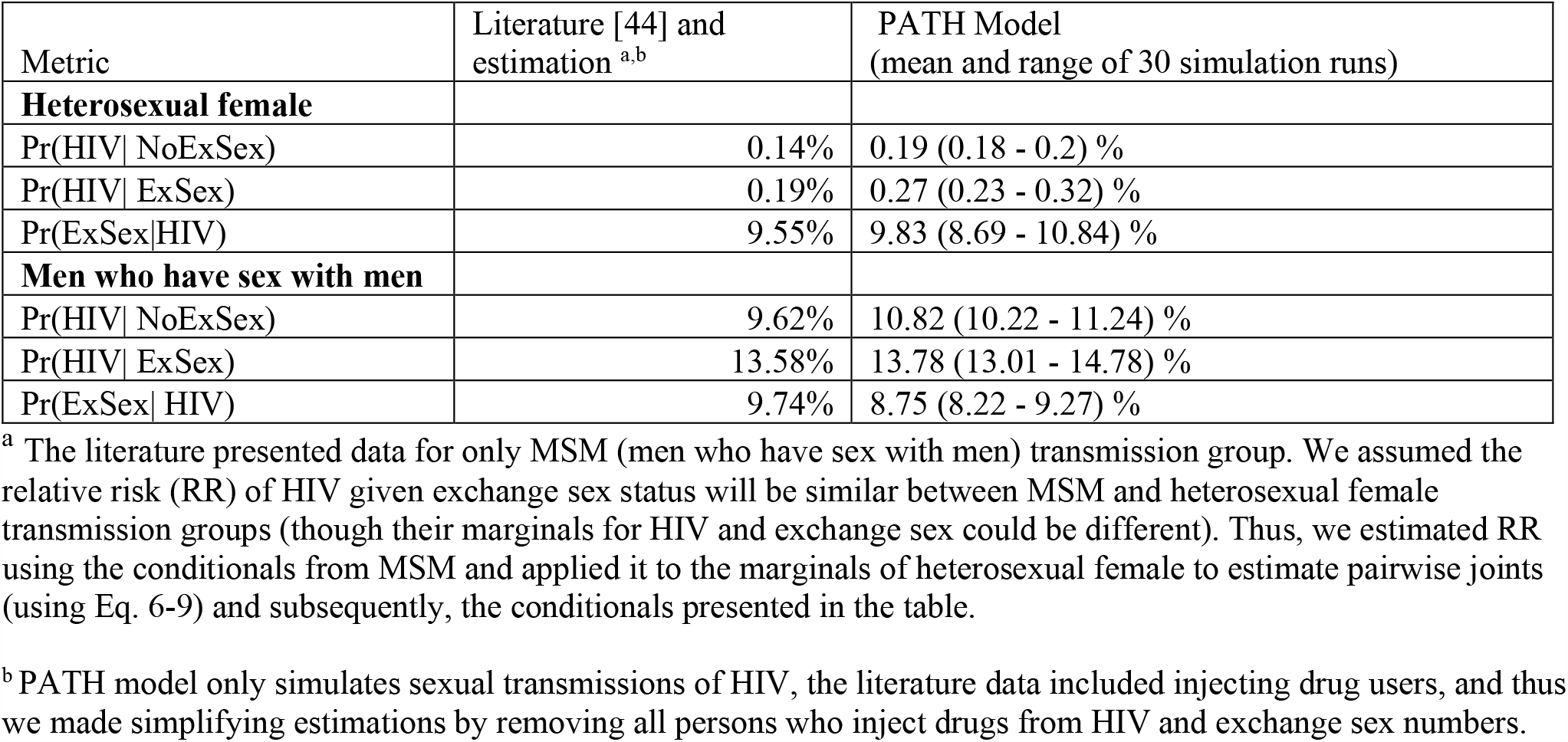
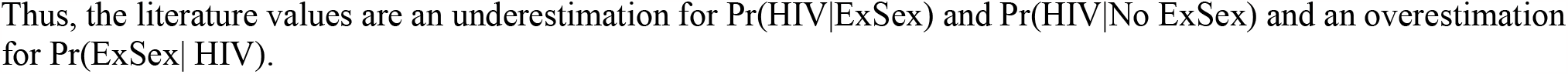
Validating PATH model simulation outcomes by comparing to literature estimates.

As expected from transmission dynamics, the prevalence of exchange sex is higher among persons with HIV than the overall population (Table 2b), with prevalence at about 9.8% among HIV infected women and 8.7% among MSM (which validates well to literature data as noted in Table 2a) compared to 7.1% among the general population. The prevalence of social conditions among PWH who exchange sex was higher than among PWH with no exchange sex, as seen by the relative risk values in Table 2b. Among HIV infected women, the relative risk values were 14.4, 69.6, 7.02, and 1.10, for Employment, Housing, Poverty, and Education, respectively. Among HIV infected MSM, the relative risk values were 3.20, 11.61, 2.94, and 1.07, for Employment, Housing, Poverty, and Education, respectively. To note here that, as exchange sex was the only mediator modeled here to link sexual behavior to social conditions, we can expect social conditions among the HIV-infected population to not be representative of reality. Thus, these results are presented to only highlight the social needs among persons who exchange sex (and not among all HIV infected persons), to serve as demonstration for potential analyses that can be conducted by modeling behaviors as functions of social conditions, especially when evaluating impact of interventions.

**Table 2b:**
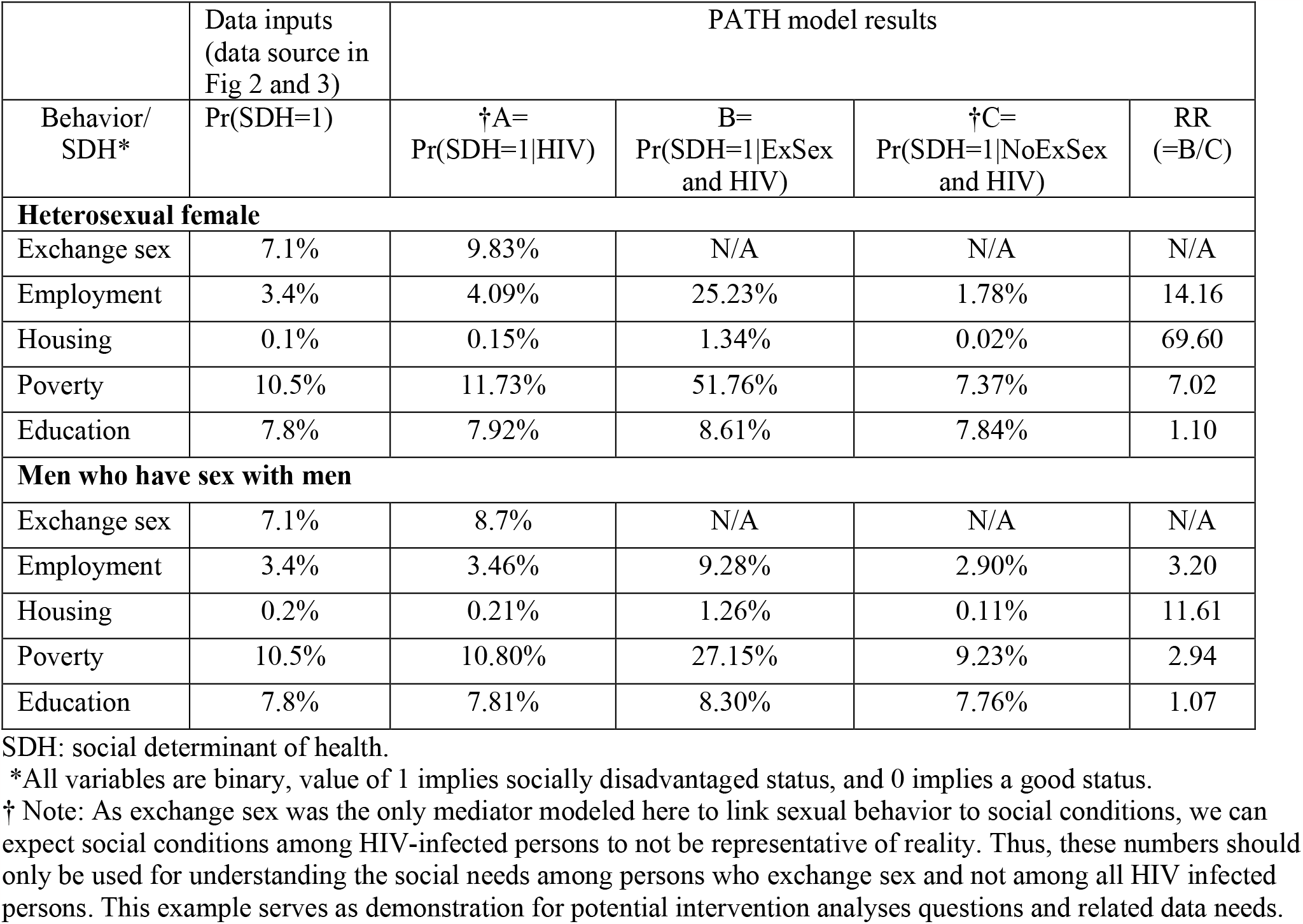
Social conditions among persons who exchange sex and are HIV-infected in the PATH simulation model.

If an intervention strategy successfully addresses the social needs of persons who exchange sex such that the number of partners and condom use among those who exchange sex is similar to those who do not, then we can expect a 10-year cumulative national incidence reduction of 6% (2.5% to 14%), the average and range across 30 simulation runs.

## 4. DISCUSSION

As social conditions are drivers of behaviors that increase risk of HIV, modeling SDH into decision-analytic mechanistic models will help evaluate structural interventions alongside pharmaceutical interventions. We present a method for building such a model. We present a Markov random field model to estimate joint distributions of social conditions and related variables. The joint distributions can be used in any mechanistic model, to add SDH and model behaviors as functions of SDH, though the mechanisms of modeling these may vary across modeling techniques. For example, compartmental models could use the joint distributions to create compartments of mutually exclusive feature combinations (e.g., degree =2, Housing =no, Poverty = yes), whereas agent-based models could use it to model individual-level features.

We also present an approach to model behaviors as functions of social conditions in mechanistic models. In numerical analyses, we applied our method to PATH 4.0, an agent-based network simulation model of HIV in the United States, to add social conditions for each agent and simulate care and sexual behaviors as functions of their social conditions. As demonstration for the potential significance of such a model, we conducted what-if analyses assuming an ideal 100% efficacious intervention strategy. Our numerical analyses suggests that, for the social conditions modeled here, if we are able to intervene on those people with the social needs, such that their retention-in-care behavior increases to become equal to that among persons with no SDH, the overall viral suppression among PWDH can increase from 65.5% to 80%, resulting in a 10-year cumulative national incidence reduction of 29% (20% to 41%). Our results also suggest that if we address the social needs modeled here among persons who exchange sex, such that their sexual behavior becomes equal to that among those who do not exchange sex, we can expect a 10-year cumulative national incidence reduction of 6% (2.5% to 14%).

Our model is subject to limitations. This work is limited to be a demonstration case study of a suitable methodology for modeling behaviors as functions of social conditions, and does not comprehensively include all social conditions. The joint distribution estimation method fits closest to uniform distribution subject to constraints, and thus the estimations are highly reliant on the features (social conditions and associations), used in the MRF model. Data on associations between variables are limited and conducted on smaller observational studies. To avoid data bias, we only used the relative risk metrics from these smaller studies and used marginal distributions that are more representative of the full population, but the caveat is that it assumes that the associations in small samples are representative of the full population. To fully account for disparities across populations, data should be specific to sub-populations, we only modeled heterogeneity by transmission group in this demonstration example, modeling data by race and ethnicity could be key features in future work. We assumed causality between behaviors and social conditions, i.e., an intervention strategy that successfully intervenes all social needs will change behaviors to be equal to those with no social conditions. Though we did not assume causality between any two social conditions, we assumed an intervention strategy will collectively address all social needs. For intervention analyses, more specific data on the effectiveness of interventions on social conditions and behaviors should be incorporated. Validation of such models can be a challenge, we only validated some of the metrics from the sexual behavioral model, highlighting consequences of data gaps in other metrics.

## 5. CONCLUSIONS

We provide an overall methodological approach for modeling social conditions into mechanistic disease prediction and intervention decision-analytic models. While the cost of intervention programs can be estimated from small controlled trials, the model will be instrumental in estimating its impact on the national disease burden, using the efficacy and costs of structural, behavioral, and biomedical interventions from those programs to model changes in social conditions and behaviors in the national context. Such a model will serve as a decision-analytic tool to evaluate alternative combinations of intervention programs, to subsequently identify intervention strategies that are the most cost-effective in reducing overall disease burden, address social needs, and reduce health disparities.

## Data Availability

All data produced in the present work are contained in the manuscript

## Funding

This work was supported by the National Science Foundation, United States, under NSF 1915481. The funding agreement ensured the authors’ independence in designing the study, interpreting the data, writing, and publishing the report.

## Authors’ contributions

CG was involved in conception of study, model development, analyses and interpretation of findings, and manuscript preparation. AK was involved in model development and analyses.

## Declaration of competing interest

CG is a Guest Researcher at the Division of HIV Prevention, Centers for Disease Control and Prevention. The findings and conclusions of this report are those of the authors and do not necessarily represent the official position of the Centers for Disease Control and Prevention. CG and AK declare no conflict of interest.

## Acknowledgements

We like to acknowledge Drs. Paul Farnham and Cynthia Lyles, from the Centers for Disease Control and Prevention and Drs. Peter Haas and Hari Balasubramanian from the University of Massachusetts Amherst for their inputs on the work.

## Notes

### Competing Interest Statement

The authors have declared no competing interest.

## References

[1] “HIV Treatment,” Division of HIV Prevention, National Center for HIV, Viral Hepatitis, STD, and TB Prevention, Centers for Disease Control and Prevention. [Online]. Available: <https://w>ww.cdc.gov/hiv/basics/livingwithhiv/treatment.html

[2] “PrEP Effectiveness,” Division of HIV Prevention, National Center for HIV, Viral Hepatitis, STD, and TB Prevention, Centers for Disease Control and Prevention. [Online]. Available: <https://w>ww.cdc.gov/hiv/basics/prep/prep-effectiveness.html

[3] “Centers for Disease Control and Prevention. Estimated HIV incidence and prevalence in the United States, 2015–2019. HIV Surveillance Supplemental Report 2021;26(No. 1). http://www.cdc.gov/hiv/library/reports/hiv-surveillance.html. Published May 2021. Accessed [11/20/201].”

[4] A. Bingham, R. K. Shrestha, N. Khurana, E. U. Jacobson, and P. G. Farnham, “Estimated Lifetime HIV-Related Medical Costs in the United States,” Sex Transm Dis, vol. 48, no. 4, pp. 299–304, Apr. 2021, doi: 10.1097/OLQ.0000000000001366.

[5] J. Craddock, A. Barman-Adhikari, K. M. Combs, A. Fulginiti, and E. Rice, “Individual and Social Network Correlates of Sexual Health Communication Among Youth Experiencing Homelessness,” AIDS Behav, vol. 24, no. 1, pp. 222–232, Jan. 2020, doi: 10.1007/s10461-019-02646-x.

[6] B. F. Henwood, H. Rhoades, B. Redline, E. Dzubur, and S. Wenzel, “Risk behaviour and access to HIV/AIDS prevention services among formerly homeless young adults living in housing programmes,” AIDS Care, vol. 32, no. 11, pp. 1457–1461, Nov. 2020, doi: 10.1080/09540121.2019.1699643.

[7] D. S. Maria, S. S. Daundasekara, D. C. Hernandez, W. Zhang, and S. C. Narendorf, “Sexual risk classes among youth experiencing homelessness: Relation to childhood adversities, current mental symptoms, substance use, and HIV testing,” PLOS ONE, vol. 15, no. 1, p. e0227331, Jan. 2020, doi: 10.1371/journal.pone.0227331.

[8] J. P. Edidin, Z. Ganim, S. J. Hunter, and N. S. Karnik, “The mental and physical health of homeless youth: a literature review,” Child Psychiatry Hum Dev, vol. 43, no. 3, pp. 354–375, Jun. 2012, doi: 10.1007/s10578-011-0270-1.

[9] “cdc-hiv-surveillance-special-report-number-25.pdf.” Accessed: Nov. 20, 2021. [Online]. Available: <https://w>ww.cdc.gov/hiv/pdf/library/reports/surveillance/cdc-hiv-surveillance-special-report-number-25.pdf

[10] Y. L. Huang et al., “Nearly Half Of US Adults Living With HIV Received Federal Disability Benefits In 2009,” Health.Aff.(Millwood), vol. 34, no. 10, pp. 1657–1665, 2015, doi: 10.1377/hlthaff.2015.0249.

[11] “The White House. 2022. National HIV/AIDS Strategy Federal Implementation Plan. Washington, DC. https://files.hiv.gov/s3fs-public/2022-09/NHAS_Federal_Implementation_Plan.pdf Last accessed 2-28-2023.”

[12] “The White House. 2021. National HIV/AIDS Strategy for the United States 2022–2025. Washington, DC. https://files.hiv.gov/s3fs-public/NHAS-2022-2025.pdf Last accesses 2-28-2023.”

[13] “Compendium of Evidence-Based Interventions and Best Practices for HIV Prevention, Centers for Disease Control and Prevention, <https://www.cdc.gov/hiv/research/interventionresearch/compendium/si/index.html >[last accessed August 2022]”.

[14] A. A. Adimora and J. D. Auerbach, “Structural interventions for HIV prevention in the United States,” J.Acquir.Immune Defic.Syndr., vol. 55, no. p S132–S135, 2010, doi: 10.1097/QAI.0b013e3181fbcb38.

[15] T. A. Sipe, T. L. Barham, W. D. Johnson, H. A. Joseph, M. L. Tungol-Ashmon, and A. O’Leary, “Structural Interventions in HIV Prevention: A Taxonomy and Descriptive Systematic Review,” AIDS.Behav., vol. 21, no. 12, pp. 3366–3430, 2017, doi: 10.1007/s10461-017-1965-5.

[16] E. E. Friedman, H. D. Dean, and W. A. Duffus, “Incorporation of Social Determinants of Health in the Peer-Reviewed Literature: A Systematic Review of Articles Authored by the National Center for HIV/AIDS, Viral Hepatitis, STD, and TB Prevention,” Public Health Rep, vol. 133, no. 4, pp. 392–412, Aug. 2018, doi: 10.1177/0033354918774788.

[17] K. M. Blankenship, S. J. Bray, and M. H. Merson, “Structural interventions in public health,” AIDS, vol. 14, no. Suppl 1:S11–21, 2000.

[18] K. M. Blankenship, S. R. Friedman, S. Dworkin, and J. E. Mantell, “Structural interventions: concepts, challenges and opportunities for research,” J.Urban Health, vol. 83, no. 1, pp. 59–72, 2006, doi: 10.1007/s11524-005-9007-4.

[19] T. R. Frieden, “A framework for public health action: the health impact pyramid,” Am.J.Public Health, vol. 100, no. 4, pp. 590–595, 2010, doi: 10.2105/AJPH.2009.185652.

[20] R. A. Bonacci and D. R. Holtgrave, “Evaluating the impact of the US National HIV/AIDS Strategy, 2010-2015,” AIDS.Behav., vol. 20, no. 7, pp. 1383–1389, 2016, doi: 10.1007/s10461-016-1416-8.

[21] D. R. Holtgrave and R. Greenwald, “A SWOT Analysis of the Updated National HIV/AIDS Strategy for the U.S., 2015-2020,” AIDS.Behav., vol. 20, no. 1, pp. 1–6, 2016, doi: 10.1007/s10461-015-1193-9.

[22] C. Gopalappa, S. L. Sansom, P. G. Farnham, and Y.-H. Chen, “Combinations of interventions to achieve a national HIV incidence reduction goal: insights from an agent-based model,” AIDS (London, England), vol. 31, no. 18, pp. 2533–2539, 2017, doi: 10.1097/QAD.0000000000001653.

[23] N. Khurana et al., “Impact of Improved HIV Care and Treatment on PrEP Effectiveness in the United States, 2016-2020,” J.Acquir.Immune Defic.Syndr., vol. 78, no. 4, pp. 399–405, 2018, doi: 10.1097/QAI.0000000000001707.

[24] S. N. Khatami, C. Gopalappa, and Mechanical and Industrial Engineering Department, University of Massachusetts Amherst, Amherst, MA 01003, USA, “A reinforcement learning model to inform optimal decision paths for HIV elimination,” MBE, vol. 18, no. 6, pp. 7666–7684, 2021, doi: 10.3934/mbe.2021380.

[25] J. W. Hogan, N. Galai, and W. W. Davis, “Modeling the Impact of Social Determinants of Health on HIV,” AIDS Behav, vol. 25, no. Suppl 2, pp. 215–224, Nov. 2021, doi: 10.1007/s10461-021-03399-2.

[26] D. Jahagirdar et al., “Incidence of HIV in Sub-Saharan Africa, 2000–2015: The Interplay Between Social Determinants and Behavioral Risk Factors,” AIDS Behav, vol. 25, no. S2, pp. 145–154, Nov. 2021, doi: 10.1007/s10461-021-03279-9.

[27] R. B. de Oliveira et al., “Incorporating social determinants of health into the mathematical modeling of HIV/AIDS,” Sci Rep, vol. 12, no. 1, p. 20541, Nov. 2022, doi: 10.1038/s41598-022-24459-0.

[28] M. C. D. Stoner et al., “Modeling Cash Plus Other Psychosocial and Structural Interventions to Prevent HIV Among Adolescent Girls and Young Women in South Africa (HPTN 068),” AIDS Behav, vol. 25, no. S2, pp. 133–143, Nov. 2021, doi: 10.1007/s10461-021-03158-3.

[29] K. Shannon et al., “Global epidemiology of HIV among female sex workers: influence of structural determinants,” Lancet, vol. 385, no. 9962, pp. 55–71, Jan. 2015, doi: 10.1016/S0140-6736(14)60931-4.

[30] D. Rasella et al., “Evaluating the impact of social determinants, conditional cash transfers and primary health care on HIV/AIDS: Study protocol of a retrospective and forecasting approach based on the data integration with a cohort of 100 million Brazilians,” PLoS ONE, vol. 17, no. 3, p. e0265253, Mar. 2022, doi: 10.1371/journal.pone.0265253.

[31] S. Singh, A. M. France, Y.-H. Chen, P. G. Farnham, A. M. Oster, and C. Gopalappa, “Progression and transmission of HIV (PATH 4.0)-A new agent-based evolving network simulation for modeling HIV transmission clusters,” Math Biosci Eng, vol. 18, no. 3, pp. 2150–2181, Mar. 2021, doi: 10.3934/mbe.2021109.

[32] K. P. Murphy, Machine learning: a probabilistic perspective. Cambridge, MA: MIT Press, 2012.

[33] “Centers for Disease Control and Prevention (CDC), Understanding the HIV care continuum, <https://w>ww.cdc.gov/hiv/pdf/library/factsheets/cdc-hiv-care-continuum.pdf. Last Accessed (2-23-2023),” 2019.

[34] “Centers for Disease Control and Prevention, Atlas Plus, <https://w>ww.cdc.gov/nchhstp/atlas/about-atlas.html]Last Accessed August 2022].”

[35] M. Eden, R. Castonguay, B. Munkhbat, H. Balasubramanian, and C. Gopalappa, “Agent-based evolving network modeling: a new simulation method for modeling low prevalence infectious diseases,” Health Care Manag Sci, vol. 24, no. 3, pp. 623–639, Sep. 2021, doi: 10.1007/s10729-021-09558-0.

[36] C. Centers for Disease and Prevention, “Vital signs: HIV testing and diagnosis among adults--United States, 2001-2009,” MMWR. Morbidity and mortality weekly report, vol. 59, no. 47, pp. 1550–5, 2010.

[37] “Centers for Disease Control and Prevention. HIV Surveillance Reports, <https://www.cdc.gov/hiv/library/reports/hiv-surveillance.html, [Last Accessed August 2022]”.

[38] D. Broz et al., “HIV infection and risk, prevention, and testing behaviors among injecting drug users -- National HIV Behavioral Surveillance System, 20 U.S. cities, 2009,” MMWR Surveill.Summ., vol. 63, no. 6, pp. 1–51, 2014.

[39] K. Buchacz et al., “CD4 cell counts at HIV diagnosis among HIV Outpatient Study participants, 2000-2009,” AIDS Res Treat, vol. 2012, p. 869841, 2012, doi: 10.1155/2012/869841.

[40] M. Reece, D. Herbenick, V. Schick, S. A. Sanders, B. Dodge, and J. D. Fortenberry, “Background and considerations on the National Survey of Sexual Health and Behavior (NSSHB) from the investigators,” J Sex Med, vol. 7 Suppl 5, pp. 243–5, 2010, doi: 10.1111/j.1743-6109.2010.02038.x.

[41] A. Chandra, W. D. Mosher, C. Copen, and C. Sionean, “Sexual behavior, sexual attraction, and sexual identity in the United States: data from the 2006-2008 National Survey of Family Growth,” Natl Health Stat Report, vol. 2011/05/13, no. 36, pp. 1–36, 2011.

[42] D. Kanny et al., “A Key Comprehensive System for Biobehavioral Surveillance of Populations Disproportionately Affected by HIV (National HIV Behavioral Surveillance): Cross-sectional Survey Study,” JMIR Public Health Surveill, vol. 8, no. 11, p. e39053, Nov. 2022, doi: 10.2196/39053.

[43] S. M. Jenness, P. Kobrak, T. Wendel, A. Neaigus, C. S. Murrill, and H. Hagan, “Patterns of exchange sex and HIV infection in high-risk heterosexual men and women,” J Urban Health, vol. 88, no. 2, pp. 329–341, Apr. 2011, doi: 10.1007/s11524-010-9534-5.

[44] L. M. Nerlander et al., “Exchange Sex and HIV Infection Among Men Who Have Sex with Men: 20 US Cities, 2011,” AIDS Behav, vol. 21, no. 8, pp. 2283–2294, Aug. 2017, doi: 10.1007/s10461-016-1450-6.

[45] “Centers for Disease Control and Prevention/ Agency for Toxic Substances and Disease Registry/ Geospatial Research, Analysis, and Services Program. CDC/ATSDR Social Vulnerability Index [Insert 2018] Database [US]. <https://www.atsdr.cdc.gov/placeandhealthsvi/data_documentation_download.html >Accessed on [1-16-2023].”

[46] R. H. Gokhale, J. Weiser, P. S. Sullivan, Q. Luo, F. Shu, and H. Bradley, “Depression Prevalence, Antidepressant Treatment Status, and Association with Sustained HIV Viral Suppression Among Adults Living with HIV in Care in the United States, 2009-2014,” AIDS Behav, vol. 23, no. 12, pp. 3452–3459, Dec. 2019, doi: 10.1007/s10461-019-02613-6.

[47] “Centers for Disease Control and Prevention. Social determinants of health among adults with diagnosed HIV infection, 2019. HIV Surveillance Supplemental Report 2022;27(No. 2). http://www.cdc.gov/hiv/library/reports/hiv-surveillance.html. Published March 2022. Accessed [July 2022].”

[48] “Centers for Disease Control and Prevention. Behavioral and Clinical Characteristics of Persons with Diagnosed HIV Infection—Medical Monitoring Project, United States, 2018 Cycle (June 2018– May 2019). HIV Surveillance Special Report 25. <https://www.cdc.gov/hiv/library/reports/hiv->surveillance.html. Published May 2020. Accessed [2-23-2023].”

[49] H. Bradley, A. H. Viall, P. M. Wortley, A. Dempsey, H. Hauck, and J. Skarbinski, “Ryan White HIV/AIDS Program Assistance and HIV Treatment Outcomes,” Clin Infect Dis, vol. 62, no. 1, pp. 90–98, Jan. 2016, doi: 10.1093/cid/civ708.

[50] C. R. Lesko, H. E. Hutton, A. T. Fojo, N. M. Shen, R. D. Moore, and G. Chander, “Depression and HIV viral nonsuppression among people engaged in HIV care in an urban clinic, 2014-2019,” AIDS, vol. 35, no. 12, pp. 2017–2024, Oct. 2021, doi: 10.1097/QAD.0000000000003005.

[51] D. Muthulingam, J. Chin, L. Hsu, S. Scheer, and S. Schwarcz, “Disparities in engagement in care and viral suppression among persons with HIV,” J Acquir Immune Defic Syndr, vol. 63, no. 1, pp. 112–119, May 2013, doi: 10.1097/QAI.0b013e3182894555.

[52] R. Stein et al., “Factors Associated with HIV Antiretroviral Therapy among Men Who Have Sex with Men in 20 US Cities, 2014,” J Urban Health, vol. 96, no. 6, pp. 868–877, Dec. 2019, doi: 10.1007/s11524-019-00386-w.

[53] D. P. Kidder, R. J. Wolitski, M. L. Campsmith, and G. V. Nakamura, “Health Status, Health Care Use, Medication Use, and Medication Adherence Among Homeless and Housed People Living With HIV/AIDS,” Am J Public Health, vol. 97, no. 12, pp. 2238–2245, Dec. 2007, doi: 10.2105/AJPH.2006.090209.

[54] D. P. Kidder, R. J. Wolitski, S. L. Pals, and M. L. Campsmith, “Housing status and HIV risk behaviors among homeless and housed persons with HIV,” J Acquir Immune Defic Syndr, vol. 49, no. 4, pp. 451–455, Dec. 2008, doi: 10.1097/qai.0b013e31818a652c.

[55] A. Aidala, J. E. Cross, R. Stall, D. Harre, and E. Sumartojo, “Housing status and HIV risk behaviors: implications for prevention and policy,” AIDS Behav, vol. 9, no. 3, pp. 251–265, Sep. 2005, doi: 10.1007/s10461-005-9000-7.

[56] “National Alliance to End Homelessness, State of Homelessness: 2022 Edition, https://endhomelessness.org/homelessness-in-america/homelessness-statistics/state-of-homelessness-2021/ [Last Accesed 1-6-2023].”

[57] “Exchange Sex Among Heterosexual Women at Risk for HIV Infection <https://www.michigan.gov/->/media/Project/Websites/mdhhs/Folder3/Folder5/Folder2/Folder105/Folder1/Folder205/Exchange_Sex_Fact_Sheet.pdf?rev=eb7f8186aaa24cc38d93c2fe265b5e37 Last Access 1-16-2023.”

